# Agentic Chart Review from Longitudinal Clinical Notes: a Lung Cancer Guideline Concordance Use Case

**DOI:** 10.64898/2026.06.02.26354727

**Authors:** Yuhang Jiang, Xing He, Xuguang Ai, Ramya Keerthi Majji, Rohan Maniar, Shadia Jalal, David A. Fedele, Jessica Hollenbach, Jinsong Liu, Yan Zhuang, Yiye Zhang, Jiang Bian

## Abstract

Clinical chart abstraction extracts structured patient variables from longitudinal clinical notes but is labor-intensive and difficult to scale. We evaluated LLM agents for question-guided chart review using lung cancer molecular testing guideline concordance as a use case. Two configurations were compared: (1) sequential note review using metadata and chronology, and (2) the same framework augmented with keyword-based note search. Gold-standard labels were established by human annotators. The search-enabled agent achieved higher accuracy (92.4% vs. 83.5%) and reduced errors by more than half (41 vs. 89) by retrieving evidence from long, heterogeneous note histories. In guideline concordance evaluation, most determinate patient–rule assessments were concordant (80.7%), while most apparent non-concordance reflected missing molecular testing documentation rather than documented care deviations. These results suggest tool-augmented LLM agents can approximate key aspects of human chart review and support scalable information extraction from longitudinal clinical documentation.

## Introduction

Chart abstraction — the systematic extraction of structured, patient-level variables from electronic health records (EHRs), especially from large collections of unstructured clinical notes — is a foundational task in clinical research, registry maintenance, and quality-of-care monitoring. In practice, it requires trained clinicians or research staff to read through potentially large volumes of longitudinal documentation, identify relevant evidence, and record a consistent set of predefined variables for each patient. This process is labor intensive, subject to inter-reviewer variability, and difficult to scale, particularly when the target variable set is large or the underlying documentation spans long periods of care across multiple specialties and note types.

For example, cancer registries routinely require abstraction of variables such as histologic subtype, disease stage, and treatment history from unstructured clinical documentation, a task that remains predominantly manual despite decades of investment in automated approaches.^1^ Similarly, quality improvement initiatives depend on chart abstraction to assess whether observed care aligns with practice guidelines. Guideline concordance assessment for lung cancer molecular testing, for instance, requires extracting variables such as diagnostic confirmation, molecular testing and biomarker results, and treatment decisions from heterogeneous documentation, including pathology reports, oncology consultations, radiology summaries, and treatment records, and evaluating the care trajectory against established recommendations.^2–4^

Recent advances in large language models (LLMs) have demonstrated near-human accuracy for extracting structured variables from clinical notes.^5^ However, most approaches operate on single notes or require concatenating all documentation into one input, limiting scalability for patients with extensive longitudinal records. Agentic LLM systems address this by planning their own review — selecting which notes to access, in what order, and when to stop — rather than processing all documentation exhaustively.^6,7^ In an agentic architecture, the language model operates within an execution loop that interleaves reasoning with tool calls (e.g., listing documents, reading notes, or searching for keywords) and iterates until sufficient evidence is gathered. This gives the agent autonomy to adapt its retrieval strategy to each question and patient record. The resulting selective retrieval resembles how experienced abstractors prioritize document types and halt once sufficient evidence is found. Yet most evaluations of LLM-based extraction use single-note inputs or synthetic cases, leaving open how well agentic systems perform when evidence is dispersed across large, heterogeneous records accumulated over years of care.

In this study, we develop a question-driven framework for agentic chart review over longitudinal clinical records and evaluate it on lung cancer molecular testing. Eighteen guideline^17^ questions, organized in a tiered dependency structure, guide the agent through the lung cancer treatment workup from diagnosis through molecular testing to therapy initiation. To understand how retrieval tool access shapes agent planning behavior and extraction quality, we compare two agent configurations: a baseline agent that navigates notes by type and date, and a search tool-augmented agent that additionally uses keyword search to locate relevant documents before reading. This comparison isolates the effect of retrieval capability on how agents navigate large, heterogeneous records — a question that has not been systematically examined in prior work. Both configurations share the same underlying language model and generation settings. Experiments were conducted on 30 lung cancer patients, with agent outputs independently reviewed by two human annotators and adjudicated to establish reference labels. Using the best-performing configuration, we conducted a downstream guideline concordance analysis to examine real-world drivers of concordant versus non-concordant care and to distinguish true clinical exceptions from documentation gaps.

This study makes three important contributions. *First*, we present a question-driven agentic framework for chart review over longitudinal clinical records, evaluated on authentic patient data with human-verified reference labels. *Second*, we examine how retrieval tool access shapes agent planning behavior and extraction quality by comparing baseline and search tool-augmented configurations on the same underlying model. *Third*, we demonstrate a downstream guideline-concordance analysis that operationalizes extracted variables for care-quality assessment, distinguishing clinical exceptions from documentation gaps.

## Related Work

### Clinical Information Extraction from EHRs

Extracting structured variables from unstructured EHR text has been an active area of clinical natural language processing (NLP) research for over a decade, progressing from rule-based and machine learning pipelines to deep learning approaches.^5,8^ Despite steady gains in extraction accuracy, challenges related to annotation cost, domain specificity, and scalability persist. A recent review of NLP for construction of clinical registries concluded that manual abstraction remains labor intensive and error prone, although automated extraction is increasingly feasible, with generative LLMs emerging as a promising but still underexplored direction.^9^ In lung cancer, relevant variables are often distributed across multiple document types, making comprehensive extraction particularly challenging.^10^

### LLM-Based Information Extraction in Oncology

The application of LLMs to oncology information extraction has expanded rapidly since 2023. A recent scoping review identified 24 studies, with lung cancer among the most frequently examined cancers; earlier work relied on fine-tuned BERT models, while prompt-based GPT approaches have increasingly been used.^11^ Recent work has shown that LLMs can extract complex biomarker documentation from EHRs; for example, Cohen et al. found that fine-tuned LLMs outperformed conventional deep learning methods for PD-L1 extraction.^12^ Broader reviews report performance approaching human-level accuracy for well-defined oncology extraction tasks.^13^

### Agentic Architectures in Healthcare

Agentic LLM systems have emerged as a paradigm for clinical tasks where models plan and execute multi-step information gathering using external tools. Unlike static prompting, agents can selectively retrieve documents, issue search queries, and determine when sufficient evidence has been collected, enabling targeted processing of large clinical records.^7^ A systematic review of AI agents in clinical medicine identified 20 implementations across diverse tasks and found that single-agent systems with tool access achieved the largest gains, with a median 53% improvement over non-agentic baselines.^6^ Parallel work shows that selective retrieval can match or outperform long-context LLM processing on clinical EHR reasoning tasks while using far fewer tokens, supporting tool-augmented agent designs over context-stuffing approaches.^14^ Evaluation frameworks such as MedAgentBench and FHIR AgentBench provide standardized settings for assessing clinical agents over patient records, though evaluation on longitudinal multi-document chart review remains limited.^15,16^

## Methods

We identified patients with lung cancer using ICD diagnosis codes from the Indiana Network for Patient Care (INPC), a statewide health information exchange that aggregates clinical data from more than 135 healthcare entities across Indiana. Clinical notes for these patients were retrieved for analysis.

### Development dataset

We selected 30 lung cancer patients from the INPC cohort and used these cases for iterative development of the agentic pipeline and detailed annotation. Across these patients, we identified 3,386 clinical notes (mean 112.9 per patient) spanning diagnostic evaluation, pathology confirmation, staging, treatment planning, systemic therapy, and longitudinal follow-up.

### Testing dataset

INPC records were linked to the IU Health Cancer Registry to obtain structured cancer characteristics, including histologic subtype and disease stage. We then restricted the cohort to patients with non-small cell lung cancer (NSCLC), advanced-stage disease (stage III–IV) at diagnosis, and availability of at least one pathology report and one oncology consultation note within 90 days of diagnosis. This yielded a testing cohort of 90 patients, including 48 adenocarcinoma, 38 squamous cell carcinoma, and 4 other NSCLC subtypes, with 37 stage III and 53 stage IV cases. The agentic pipeline developed on the 30-patient development set was subsequently applied to this cohort for guideline concordance evaluation. This study was approved by the Indiana University Institutional Review Board (IRB protocol #30807).

### Question Framework for Chart Abstraction

We developed a fixed set of 18 clinically grounded questions derived from NCCN clinical practice guidelines for molecular testing and treatment pathways in NSCLC. The questions reflect variables commonly abstracted during clinical chart review when assessing diagnostic workup, biomarker testing, and treatment readiness. They are organized into a five-tier dependency structure, in which variables established in earlier tiers provide the clinical context required for downstream questions (**Figure 1**). The framework spans six categories: eligibility; molecular & biomarker testing; testing timing; testing results; treatment alignment; and testing completeness, capturing key elements of lung cancer diagnostic and treatment pathway such as histologic subtype, disease stage, and smoking history; molecular and biomarker testing status, modality, and timing; specific molecular findings and actionable alterations; as well as treatment alignment, turnaround time, and biomarker panel completeness. The underlying note corpus comprises diverse document types reflecting real-world practice, including general progress notes, discharge summaries, prescription records, telephone encounter notes, and oncology outpatient progress notes.

**Figure 1.**
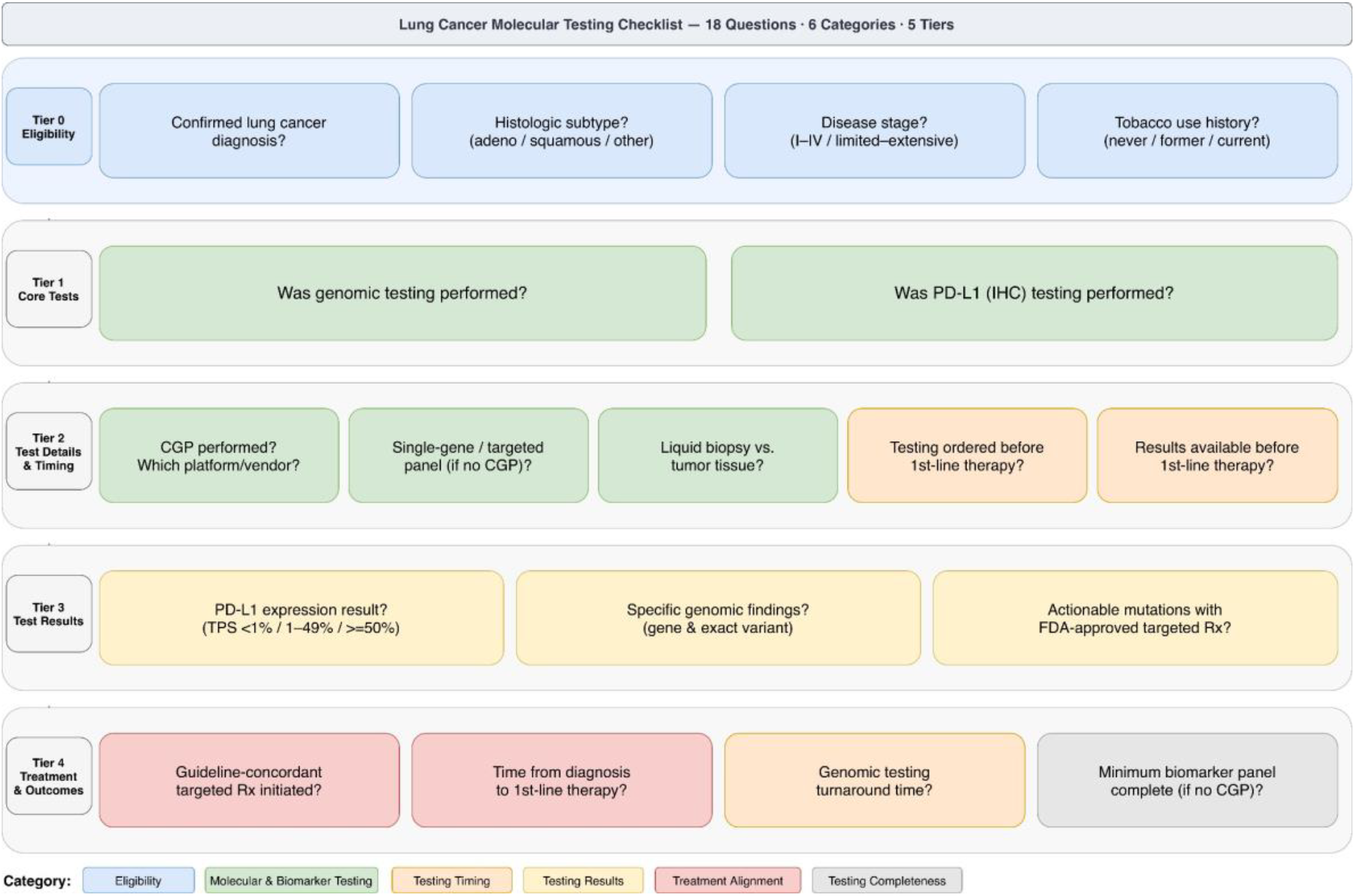
Tiered question framework for lung cancer molecular testing chart review. 18 guideline-derived questions are organized across five tiers of increasing clinical complexity and six categories. Arrows indicate dependency: downstream tiers depend on variables established in earlier tiers.

### Annotation Dataset and Label Schema

For each patient–question instance, the agent first generates a structured response comprising a direct answer, supporting evidence extracted from the notes, and a confidence rating. These outputs are then reviewed by two human annotators who independently evaluate the agent’s response against the underlying note evidence and assign a structured label. Depending on the question, labels may be binary (yes/no/unclear), categorical (e.g., smoking status, testing platform), or extraction-oriented (normalized free-text detail). Discrepancies between annotators are resolved through structured adjudication, producing final reference labels for downstream analysis.

We developed an agentic chart review workflow in which a reviewer agent answers each clinical question by navigating a patient’s note corpus with assigned tools. We compared two configurations with the same language model and generation settings but different tool access: a baseline agent that lists and reads notes by type and date, and a search-augmented agent with an additional keyword search tool. Each question runs as an independent agent session, though questions within the same tier can reuse previously summarized evidence. Execution logs and traces are retained for error analysis.

### Agent Designs

Both agents are implemented using LangChain Deep Agents (i.e., a framework supporting multi-step planning, structured tool use, and iterative execution loops) and rely on a shared Azure-hosted language model backend (**Figures 2-3**). We use GPT-5.2 deployed in the Azure OpenAI Data Zone Standard environment to ensure secure processing of patient data within the approved clinical computing environment. All generation parameters are held constant across configurations to ensure a fair comparison.

**Figure 2.**
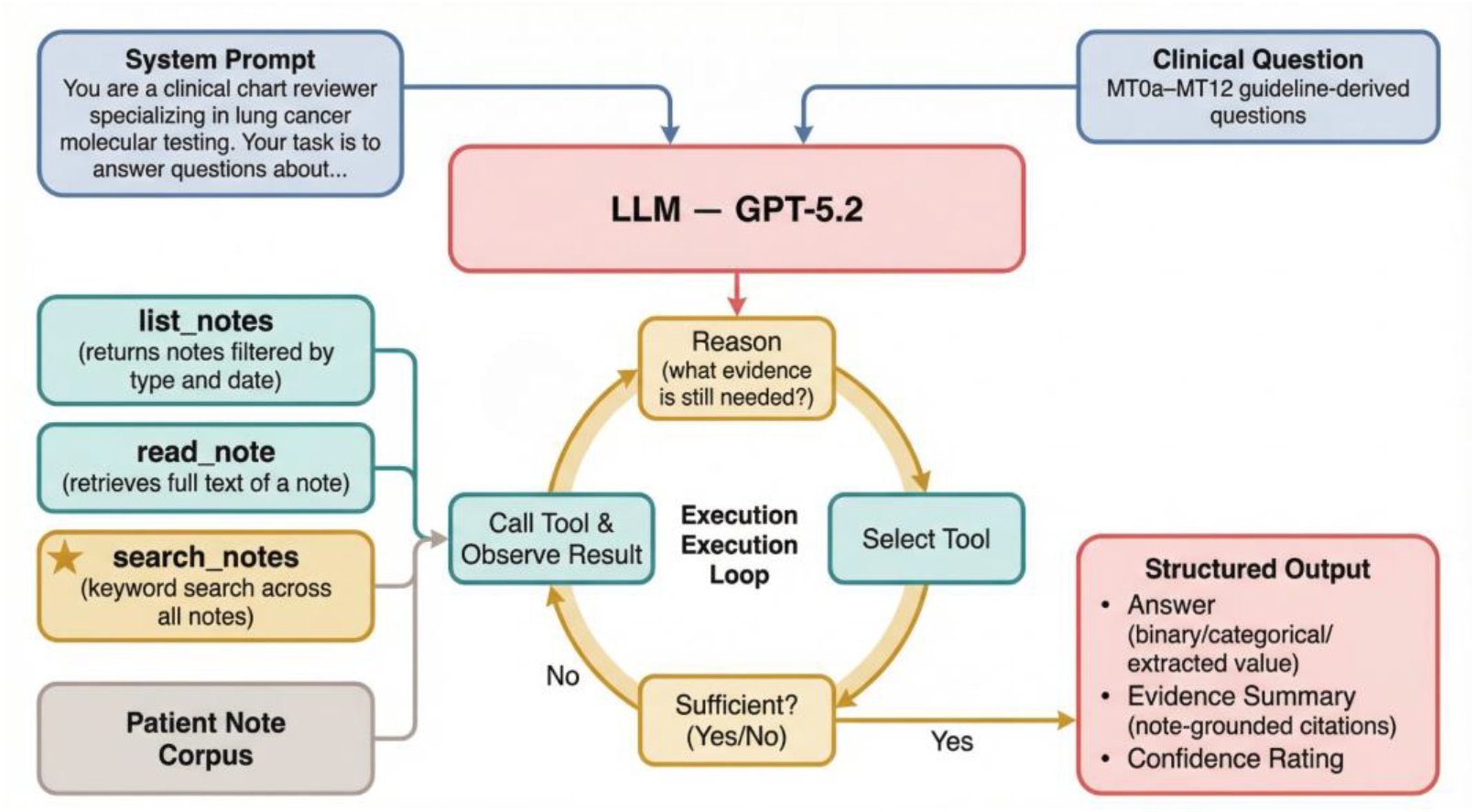
Agentic chart review architecture. The language model (GPT-5.2) operates within an iterative execution loop: at each step it reasons about what evidence is needed, calls one of the available tools (‘list_notes’, ‘read_note’, or ‘search_notes’), observes the result, and decides whether to continue gathering evidence or produce a final answer. The prompt supplies the clinical question, expected answer format, and reviewer role instructions. The output is a structured response comprising a direct answer, note-grounded evidence summary, and confidence rating.

**Figure 3.**
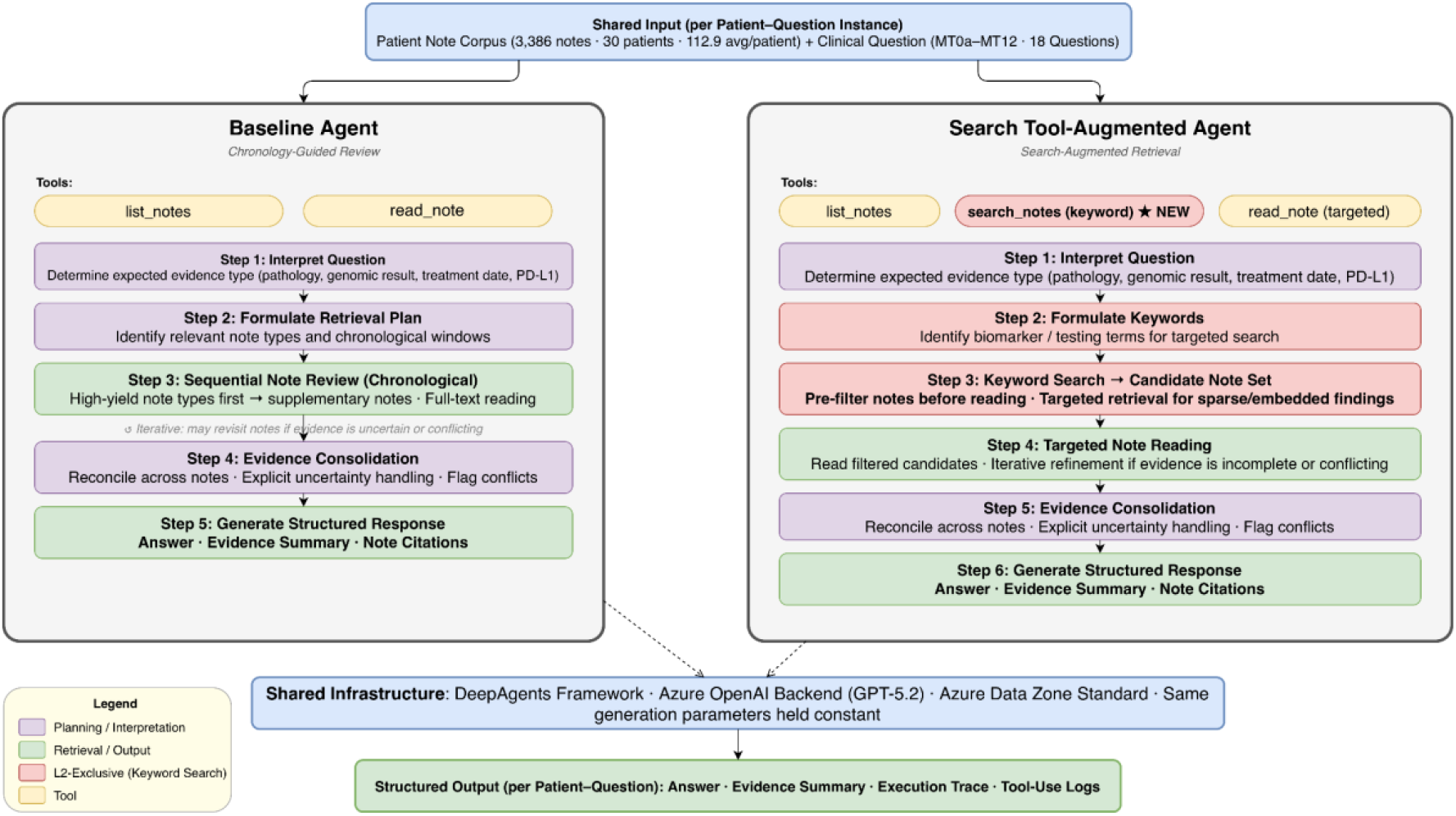
Step-by-step review workflow for each agent configuration. The baseline agent (**left**) navigates notes sequentially by type and date. The search tool-augmented agent (**right**) adds a keyword search step that pre-filters candidate notes before targeted reading. Both workflows share the same input, infrastructure, and output format (see Figure 2).

### Role and Prompting

Each agent is prompted as a clinical chart reviewer. For each patient–question instance, the agent receives: (1) the clinical question with its definition and expected answer format, (2) a system instruction defining the reviewer role and output requirements, and (3) access to the patient’s note corpus through its assigned tools. Questions are presented one at a time. The agents are prompted in zero-shot settings with no examples provided.

### Tools

The agents have access to a deliberately limited tool set. Both share two base tools: ‘list_notes’, which returns available notes filtered by type and date range, and ‘read_note’, which retrieves the full text of a specific note. The search tool-augmented agent has a third tool: ‘search_notes’, a keyword search function that queries across all notes and returns matching excerpts with note identifiers. No other tools (e.g., for patient demographics or structured data) are provided; the agents must extract information from unstructured note text.

### Reasoning and Execution Loop

As illustrated in **Figure 2**, the agent operates within an iterative loop: at each step, it reasons about what evidence is still needed, selects and calls a tool, incorporates the returned information, and decides whether to continue gathering evidence or produce a final answer. The maximum number of iterations is set to 25 without token budget or timeout limitations. The search agent selects which notes to read based on search results rather than reading all notes sequentially. The number and choice of tool calls is determined by the LLM at each step, so retrieval depth varies across questions and patients.

### Structured Output

For each question, the agent produces a structured response comprising: (a) a direct answer in the expected format (binary, categorical, or extracted value), (b) an evidence summary citing specific notes and passages that support the answer, and (c) a confidence rating. The agents are prompted to produce output in JSON format.

### Baseline vs. Search Tool-Augmented

The baseline agent can only discover relevant notes by listing and reading them sequentially, guided by note type and chronological order. The search tool-augmented agent can additionally issue keyword queries to pre-filter candidate notes before reading, enabling more targeted retrieval. This is particularly useful for locating sparse or deeply embedded findings such as biomarker results, genomic testing events, and treatment initiation decisions.

## Results

The search tool-augmented agent achieved 92.4% overall accuracy (499/540) compared with 83.5% (451/540) for the baseline (**Table 1**). The largest gains occurred on questions requiring targeted retrieval from long note histories — for example, MT1 (molecular/biomarker testing performed) improved from 70.0% to 96.7%, and MT7 (PD-L1 testing performed) from 83.3% to 100%. Some questions remained challenging for both agents, notably MT4 (tissue vs. liquid biopsy source), where accuracy was 30.0% and 40.0% respectively. F1 scores on clinically actionable positives confirm this pattern, with the search tool-augmented agent achieving a micro-averaged F1 of 0.909 versus 0.766 for the baseline. MT9 and MT12 F1 scores are not reported because the 30-patient cohort contains no positive adjudicated cases for these questions. **Table 2** summarizes the demographic and cancer characteristics of the two study cohorts: the 30-patient adjudicated cohort used for gold-standard evaluation and the 90-patient testing cohort for downstream guideline concordance assessment.

**Table 1.** Per-question extraction performance of the baseline and search tool-augmented agents on 30 lung cancer patients (540 patient–question instances). Accuracy reports correct predictions/30; F1 is computed on clinically actionable (non-negative) answers. Dashes indicate questions with no positive adjudicated cases.

| Tier | Question | Description | Baseline Acc | Baseline F1 | Search+ Acc | Search+ F1 |
| --- | --- | --- | --- | --- | --- | --- |
| 0 | MT0a | Lung cancer confirmed | 29/30 (0.967) | 0.983 | 30/30 (1.000) | 1.000 |
| 0 | MT0b | Histologic subtype | 25/30 (0.833) | 0.847 | 29/30 (0.967) | 0.967 |
| 0 | MT0c | Disease stage | 20/30 (0.667) | 0.731 | 26/30 (0.867) | 0.926 |
| 0 | MT0d | Tobacco use | 29/30 (0.967) | 0.967 | 30/30 (1.000) | 1.000 |
| 1 | MT1 | Molecular testing performed | 21/30 (0.700) | 0.714 | 29/30 (0.967) | 1.000 |
| 1 | MT7 | PD-L1 testing performed | 25/30 (0.833) | 0.737 | 30/30 (1.000) | 1.000 |
| 2 | MT2 | CGP performed/platform | 22/30 (0.733) | 0.333 | 24/30 (0.800) | 0.625 |
| 2 | MT3 | Single-gene/targeted panel | 27/30 (0.900) | 0.824 | 29/30 (0.967) | 0.947 |
| 2 | MT4 | Tissue vs. liquid biopsy | 9/30 (0.300) | 0.359 | 12/30 (0.400) | 0.500 |
| 2 | MT5 | Testing ordered before 1st-line Rx | 25/30 (0.833) | 0.667 | 29/30 (0.967) | 0.952 |
| 2 | MT6 | Results available before 1st-line Rx | 24/30 (0.800) | 0.500 | 29/30 (0.967) | 0.941 |
| 3 | MT7a | PD-L1 expression level | 27/30 (0.900) | 0.824 | 30/30 (1.000) | 1.000 |
| 3 | MT8 | Genomic findings | 26/30 (0.867) | 0.000 | 28/30 (0.933) | 0.667 |
| 3 | MT8a | Actionable mutations | 29/30 (0.967) | 0.667 | 30/30 (1.000) | 1.000 |
| 4 | MT9 | Guideline-concordant targeted Rx | 29/30 (0.967) | — | 29/30 (0.967) | — |
| 4 | MT10 | Time to 1st-line Rx | 27/30 (0.900) | 0.889 | 28/30 (0.933) | 0.897 |
| 4 | MT11 | Genomic testing turnaround | 28/30 (0.933) | 0.600 | 29/30 (0.967) | 0.727 |
| 4 | MT12 | Min biomarker panel complete | 29/30 (0.967) | — | 28/30 (0.933) | — |
|  | **Overall** |  | **451/540 (83.5%)** | **0.766** | **499/540 (92.4%)** | **0.909** |

**Table 2.** Demographic and cancer characteristics of the 30-patient development cohort and the 90-patient testing cohort. The demographic distribution of the cohort is broadly consistent with the expected clinical population of lung cancer patients, with the majority of patients aged ≥65 and NSCLC histologies predominating.

| Variable | Category | 30-patient cohort (N=30) | 90-patient cohort (N=90) |
| --- | --- | --- | --- |
| Age group | <50 | 1 (3%) | 3 (3%) |
|  | 50-64 | 12 (40%) | 27 (30%) |
| | $\geq 65$ | 17 (57%) | 60 (67%) |
| Gender | Female | 18 (60%) | 32 (36%) |
|  | Male | 12 (40%) | 58 (64%) |
| Race | White | 21 (70%) | 70 (78%) |
|  | Black | 1 (3%) | 6 (7%) |
|  | Other/Unknown | 8 (27%) | 14 (16%) |
| Ethnicity | Hispanic or Latino | 4 (13%) | 13 (14%) |
|  | Not Hispanic or Latino | 25 (83%) | 77 (86%) |
|  | Missing | 1 (3%) | 0 (0%) |
| Cancer type | Adenocarcinoma | 14 (47%) | 48 (53%) |
|  | Squamous cell carcinoma | 10 (33%) | 38 (42%) |
|  | Other NSCLC | 1 (3%) | 4 (4%) |
|  | SCLC/Neuroendocrine | 5 (17%) | 0 (0%) |
| Stage at diagnosis | I-II | 9 (30%) | 0 (0%) |
|  | III | 9 (30%) | 37 (41%) |
|  | IV | 9 (30%) | 53 (59%) |
|  | Unknown/Other | 3 (10%) | 0 (0%) |

## Discussion

### Error Analysis

We categorize errors using a LLM from agent reasoning traces into two types: (1) locate errors, where the agent fails to retrieve relevant evidence, and (2) reason errors, where evidence is retrieved but incorrectly interpreted (**Figure 4**). Both types are more frequent in the baseline agent, with locate errors showing the largest difference.

**Figure 4.**
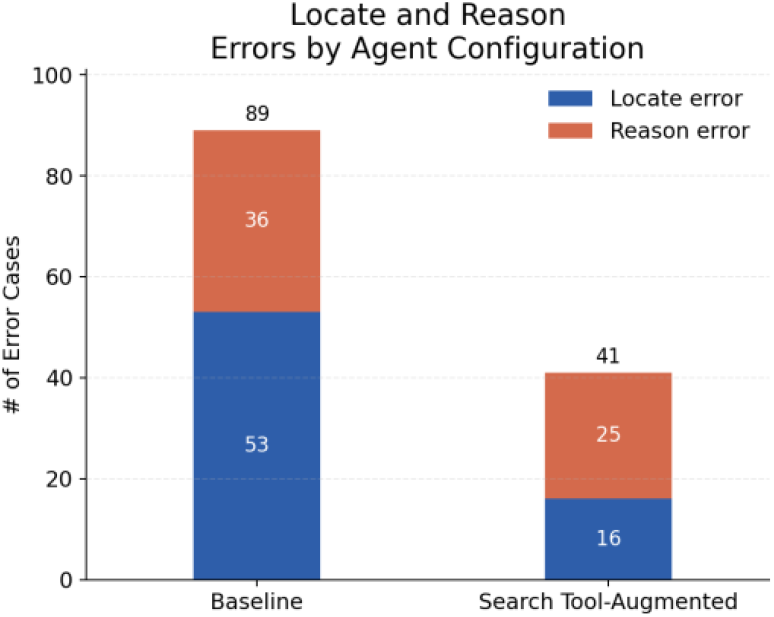
Locate and reason errors for the baseline and search-augmented agents across 540 instances.

The dominant failure pattern for the baseline agent was premature termination: the agent identified a plausible narrative from early notes and finalized the answer without checking later documentation. This was most pronounced for MT1 (molecular testing performed), where the baseline agent concluded testing was absent despite confirmatory results in subsequent notes — a pattern analogous to the “*satisfaction of search*” error in human chart review.

The search tool-augmented agent reduced both locate and reason errors compared with the baseline agent, but two residual reasoning patterns persisted. First, retrieval fixation: search surfaced targeted snippets, but the agent over-relied on a single result while missing nearby contextual constraints. Second, date linkage errors on interval questions (e.g., MT10, Time to 1st-line Rx), where diagnosis date and treatment date were incorrectly paired.

Here we provide one concrete example: the question asked for the interval between the initial lung cancer diagnosis and the initiation of first-line systemic therapy. The chart contained two potential diagnosis anchors. In the 09-25-2020 oncology note, the record referenced “*Pathology Report 9/22/2020*” and later summarized the same workup as “*Cytology 09/22/2020 … POSITIVE FOR MALIGNANCY*,” followed by “*Right Lung Cancer; Hilar Region*” and an order to begin chemotherapy in one week. In contrast, the 10-02-2020 oncology note listed “*Diagnosis Date: 10/1/2020*” and documented “*start tx today with carbo, taxotere and keytruda every 3 weeks*.”

Although the search agent retrieved the relevant notes, it anchored the interval to the later diagnosis date and predicted a time-to-treatment of 1 day (10/01/2020–10/02/2020). The gold interval was 10 days (09/22/2020–10/02/2020), indicating a reasoning error rather than a retrieval failure.

### Token Cost Analysis

The search tool–augmented agent used more tokens per patient than the baseline agent due to additional retrieval (**Figure 5**). Average tokens across 18 questions were 299.9K for the baseline agent and 534.5K for the search-enabled agent, mainly from increased input tokens. Despite higher usage, the search-enabled agent reduced total errors by more than half (41 vs. 89), indicating a favorable trade-off. Token increases were greatest for questions requiring evidence synthesis across multiple notes.

**Figure 5.**
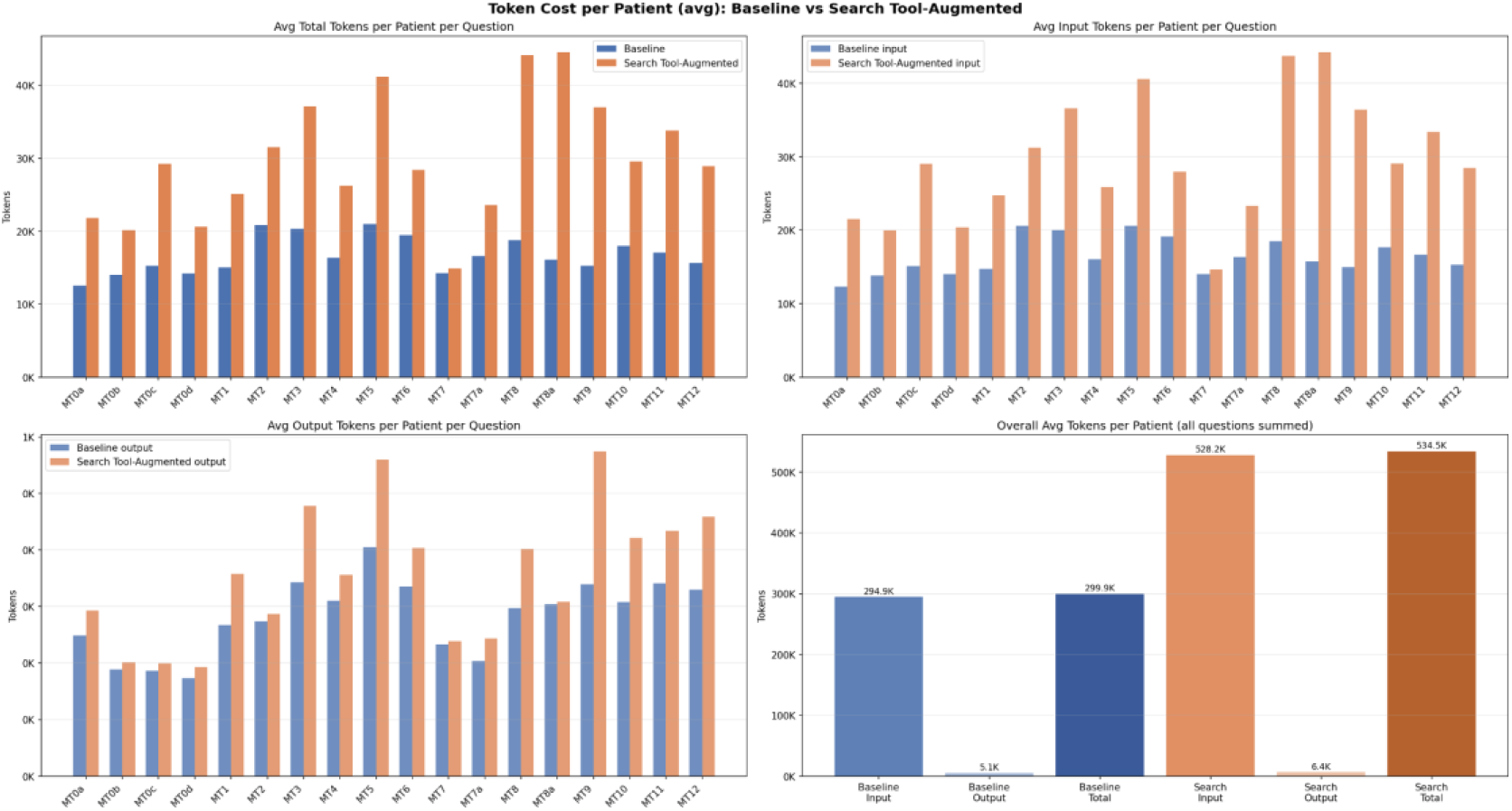
Token usage per patient for the baseline versus search tool-augmented agent across 18 chart-review questions. **Top-left**: average total tokens per patient-question; **top-right:** average input tokens; **bottom-left:** average output tokens; **bottom-right:** overall average tokens per patient (summed across all questions).

### Concordance Evaluation

To demonstrate a downstream application of the agentic extraction pipeline, we conducted a guideline concordance analysis on the 90-patient testing cohort in the lung cancer molecular testing treatment pathway (see **Methods** for cohort criteria) using outputs from the search-augmented agent. A separate LLM judge evaluated concordance based on agent-extracted evidence, including derived facts and answer texts with supporting quotes from the 18 questions.

The judge evaluated six concordance rules (**Table 3**), assigning one of three verdicts: *concordant, non-concordant*, or *uncertain* (evidence insufficient to judge). For each rule, the judge also produced a chart-grounded explanation with structured attribution for non-concordance or uncertainty.

**Table 3.**
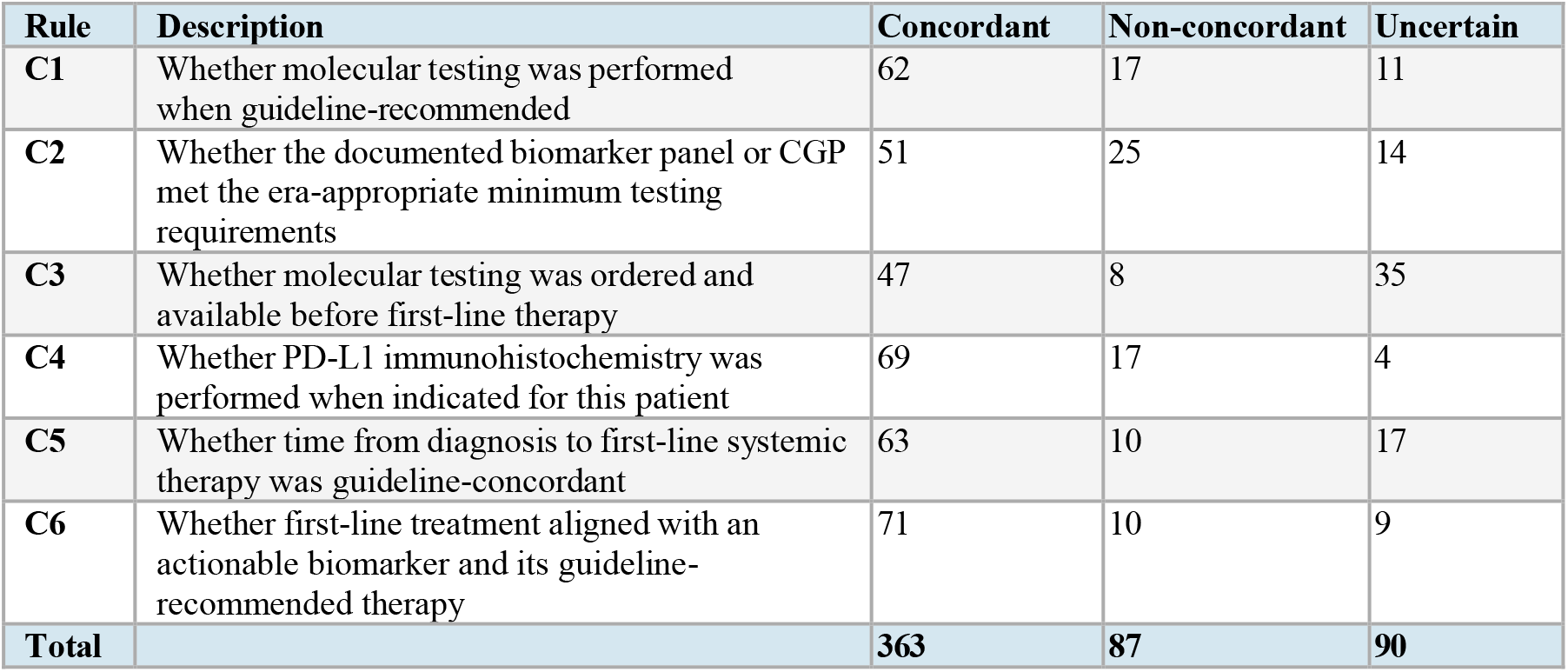
Concordance rules derived from NCCN guidelines for lung cancer molecular testing and treatment pathways. Each rule maps to one or more upstream extraction questions (MT0–MT12) and is evaluated independently per patient.

### Overall Concordance Results

Across 90 patients and 6 rules, the judge produced 540 patient–rule judgments. Of these, 363 (67.2%) were *concordant*, 87 (16.1%) were *non-concordant*, and 90 (16.7%) were *uncertain*. Among determinate judgments (excluding *uncertain*), the concordance rate was 80.7% (363/450). The *uncertain* rate (16.7%) is itself informative: it quantifies the gap between what guideline concordance assessment requires and what real-world clinical documentation provides. Out of 90 patients, 19 (21.1%) satisfy all concordance rules (their treatment pathway is concordant). 57 (63.3%) have at least one *non-concordant* rule. 14 (15.6%) have no *non-concordant* rules but have at least one *uncertain* rule.

### Reasons for non-concordance

For non-concordant cases, the judge assigned a reason (**Table 4**). The most common was missing testing documentation (49/87, 56.3%), suggesting many apparent *non-concordant* cases reflected incomplete records rather than care deviations. Other categories included planned but incomplete testing (10/87, 11.5%), treatment started before results were available (10/87, 11.5%), unexplained treatment choice or delay (10/87, 11.5%), and incorrect first-line treatment for a known biomarker (8/87, 9.2%).

**Table 4.** Reasons assigned to non-concordant cases in the 90-patient cohort after concordance adjudication.

| Reasons | Definition | Count |
| --- | --- | --- |
| *Missing testing documentation* | when required testing/results are absent from the available chart. | 49 |
| *planned but incomplete testing* | when testing was planned or partially completed but not fully resulted. | 10 |
| *treatment started before testing was ready* | when therapy began before testing was ordered/completed/resulted. | 10 |
| *wrong first-line treatment for known biomarker* | when a documented actionable biomarker did not drive first-line therapy choice. | 8 |
| *unexplained treatment choice or delay* | when non-concordance is evident but the chart does not support a more specific cause. | 10 |
| **Total** |  | **87** |

### Illustrative Cases

We present representative cases to illustrate how the agent evaluates guideline concordance.

### Correct

For the concordance rule (Testing Timing — Results):

*Were molecular testing results available before first-line therapy was initiated?*

*Compare the genomic test result date with the first-line therapy start date. If therapy began before results were available, it may indicate empiric treatment without molecular guidance. If testing was not required for the patient, mark CONCORDANT*.

The rule requires molecular testing results to be available before first-line therapy. In this case, chemotherapy began in early–mid May 2016, while the genomic report was issued on 7/15/2016. Because results were not available when treatment started, the case is classified as non-concordant.

### Error

For the concordance rule (Treatment-Mutation Concordance):

*If an actionable mutation with an FDA-approved targeted therapy was identified, was the guideline-recommended targeted therapy initiated as first-line treatment?*

*Actionable mutations with FDA-approved targeted therapies (as of 2024): ALK fusions* → *alectinib or lorlatinib; ROS1 fusions* → *crizotinib or entrectinib; KRAS G12C* → *sotorasib or adagrasib…*

The concordance rules apply current NCCN guideline recommendations to a cohort spanning 2015–2025. For treatment-alignment rules, this can produce anachronistic judgments: a 2015 patient with KRAS G12C was flagged for not receiving targeted therapy, although the first KRAS G12C inhibitor (sotorasib) was not FDA-approved until 2021.

## Conclusion

We presented an agentic framework for question-driven chart review over longitudinal clinical notes and evaluated how retrieval tool access shapes agent behavior. The search tool-augmented agent achieved 92.4% accuracy versus 83.5% for the baseline agent, with total errors reduced by more than half (41 vs. 89). Error analysis showed that baseline failures were mainly due to premature termination during sequential note reading, while remaining errors in the search-augmented agent involved retrieval fixation and temporal reasoning across notes. In downstream guideline concordance evaluation, most determinate patient–rule assessments were concordant (80.7%), and most apparent non-concordance reflected missing molecular testing documentation rather than documented care deviations. This study has several limitations, including a small annotated cohort (30 patients), reliance on a single LLM, and evaluation on one clinical domain. Future work should assess generalizability across institutions, note types, and disease areas, and explore cost-reduction strategies such as adaptive retrieval depth. These findings suggest that tool-augmented LLM agents can approximate key aspects of human chart review and support scalable information extraction from clinical records.

## Data Availability

The data used in this study are not publicly available due to patient privacy protections and institutional data use restrictions.

## Notes

### Competing Interest Statement

The authors have declared no competing interest.

### Author Declarations

Ethical approval for this work was granted by the Indiana University Institutional Review Board (IRB protocol #30807).

